# Persistence of neutralizing antibodies a year after SARS-CoV-2 infection

**DOI:** 10.1101/2021.07.13.21260426

**Authors:** Anu Haveri, Nina Ekström, Anna Solastie, Camilla Virta, Pamela Österlund, Elina Isosaari, Hanna Nohynek, Arto A. Palmu, Merit Melin

**Author notes:** Corresponding authors: Anu Haveri,; Nina Ekström. equal contribution (Anu Haveri, Nina Ekström).

## Abstract

Understanding for how long antibodies persist following Severe acute respiratory coronavirus 2 (SARS-CoV-2) infection provides important insight into estimating the duration of immunity induced by infection.

We assessed the persistence of serum antibodies following wild-type SARS-CoV-2 infection six and twelve months after diagnosis in 367 individuals of whom 13% had severe disease requiring hospitalization. We determined the SARS-CoV-2 spike (S-IgG) and nucleoprotein IgG concentrations and the proportion of subjects with neutralizing antibodies (NAb). We also measured the NAb titers among a smaller subset of participants (n=78) against a wild-type virus (B.1) and three variants of concern (VOCs): Alpha (B.1.1.7), Beta (B.1.351) and Delta (B.1.617.2).

We found that NAb against the wild-type virus and S-IgG persisted in 89% and 97% of subjects for at least twelve months after infection, respectively. IgG and NAb levels were higher after severe infection. NAb titers were significantly lower against variants compared to the wild-type virus.

## Introduction

Infection with Severe acute respiratory coronavirus 2 (SARS-CoV-2) induces antibodies in most subjects to viral nucleoprotein (N) and spike glycoprotein (S) (1). Neutralizing antibodies (NAb) against SARS-CoV-2 target the receptor-binding domain (RBD) of the spike protein and sterically interfere with the binding of the viral spike protein and the host’s angiotensin-converting enzyme 2 (ACE2) (2, 3). NAb levels are highly predictive of protection against infection and clinical disease (4) and detectable NAb have been reported to persist in most subjects at least six to twelve months after infection (5-12).

SARS-CoV-2 virus is constantly mutating yet most changes have little or no impact on its virulence (13). However, some changes are causing concerns regarding disease severity, viral transmissibility and potential escape from natural and vaccine-induced immunity (14). The World Health organization (WHO) in collaboration with international network of experts has characterized Variants of Interest (VOI) and Variants of Concern (VOC) (https://www.who.int/en/activities/tracking-SARS-CoV-2-variants/). Reduced NAb levels as compared to the wild-type virus have been shown against VOCs, especially against Beta variant, both after vaccination (12, 15-17) and nine (12) and twelve months (11) after infection. Similar reduction in NAb titers has also been reported against Delta variant from convalescent sera collected three to twelve months post symptoms or after vaccination (18, 19).

Previous infection with SARS-CoV-2 has shown to induce effective immunity and protection against reinfections in most individuals (20, 21). In animal studies, a protective antibody titer against SARS-CoV-2 infection has been suggested to be low (22, 23). Higher IgG antibody levels against SARS-CoV-2 among health care workers within three months after vaccination were found to be associated with lower infectivity (24). However, a protective threshold for humans is still under debate and subject to standardization of serological methods. The accumulating research data on the persistence of antibodies after natural infection, and neutralizing antibodies in particular, will provide important insight into estimating for how long antibodies induced by COVID-19 vaccination can be expected to persist and provide protection against emerging SARS-CoV-2 variants. In this study, we investigated the antibody persistence up to 14 months after natural SARS-CoV-2 infection and assessed the potential cross-protection by comparing the NAb levels of wild-type virus (B.1 lineage) to three VOC strains Alpha (B.1.1.7), Beta (B.1.351) and Delta (B.1.617.2).

## Results

### Study cohort characteristics

We invited a total of 2586 subjects who had experienced a PCR-confirmed SARS-CoV-2 infection six months earlier (between February 29^th^ and April 30^th^ 2020) to participate in the study. A total of 1292 (50%) participated and the median time from diagnosis to sampling was 7.6 months (range 5.9 to 9.9 months). The median age of the participants at the time of diagnosis was 50.0 years (range 17.3 to 94.3 years) and 40% were male. The majority of the participants had experienced a mild disease and 15% had been hospitalized because of COVID-19. By May 21^st^ 2021, up to 77% (995/1292) of those who participated at six months returned for a twelve month follow-up visit with a median time from diagnosis to sampling being 12.7 months (range 11.7 to 14.3 months) (Figure 1). Participant demographics and clinical characteristics are presented in Table 1.

**Figure 1.**
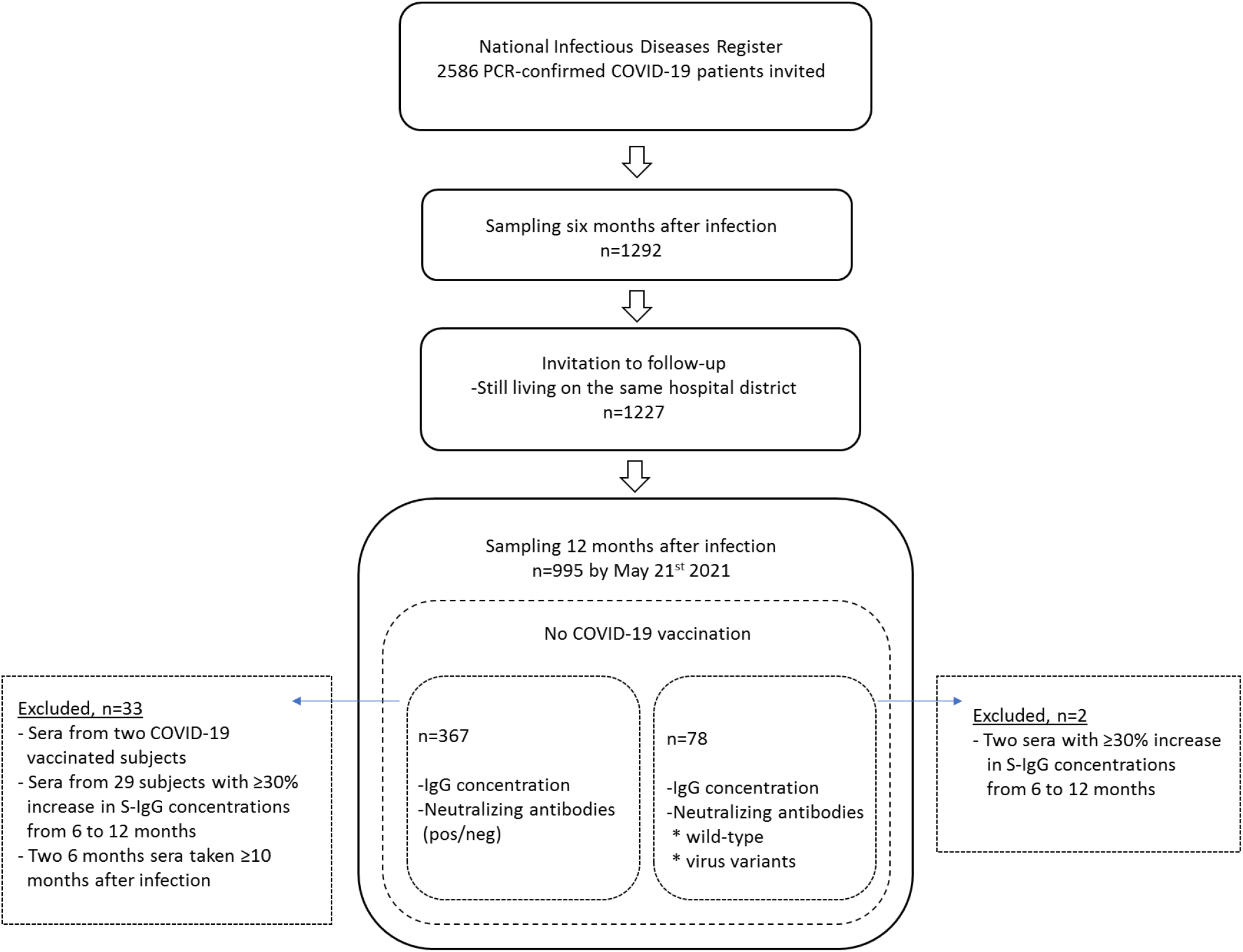
Study flow chart

**Table 1.** Demographics and clinical characteristics of study participants in the study cohorts at six and twelve months after infection.

|  | Six months participants | Twelve months participants | Study Cohort | Sub Cohort |
| --- | --- | --- | --- | --- |
| **N** |  |  |  |  |
| Six months | 1292 | - | 367 | - |
| Twelve months | - | 995 | 367 | 78 |
| **Gender** |  |  |  |  |
| Male n (%) | 520 (40%) | 386 (39%) | 159 (43%) | 40 (51%) |
| Female n (%) | 772 (60%) | 609 (61%) | 208 (57%) | 38 (49%) |
| **Age at diagnosis (median, range)** |  |  |  |  |
| <60y | 45.1 (17.3-59.9) | 47.5(17.6-59.9) | 45.9 (17.7-59.9) | 51.6 (19.0-59.7) |
| >60y | 65.1 (60.0-94.5) | 65.4 (60.0-95.6) | 63.3 (60.0-79.0) | 63.0 (60.0-81.3) |
| All | 50.0 (17.3-94.3) | 52.5 (17.6-95.6) | 48.8 (17.7-79.0) | 59.4 (19.0-81.3) |
| **Time (mo) after diagnosis at sampling** |  |  |  |  |
| Six months | 7.6 (5.9-9.9) | - | 7.6 (6.1-9.7) | - |
| Twelve months | - | 12.7 (11.7-14.3) | 12.7 (11.9-14.0) | 13.0 (12.2-13.6) |
| **Disease severity** |  |  |  |  |
| Severe | 190 (15%) | 149 (15%) | 47 (13%) | 39 (50%) |
| Mild | 1102 (85%) | 846 (85%) | 320 (87%) | 39 (50%) |

### Persistence and kinetics of SARS-CoV-2 antibodies

We first assessed the persistence of NAb and serum IgG antibodies specific to SARS-CoV-2 full-length spike protein (SFL-IgG), receptor binding domain of spike protein (RBD-IgG) and nucleoprotein (N-IgG) at six months following SARS-CoV-2 infection. We found that 89% (1148/1292) of the subjects had NAb against the wild-type virus, 96% (1240/1292) had antibodies to SFL and RBD (S-IgG) and 66% (846/1292) had N-IgG. We randomly selected to further analysis 400 subjects of the 652 subjects who had not received a SARS-CoV-2 vaccination of the 995 subjects who participated at both time points. Of the 400 subjects 33 were later excluded from the analysis due to rise in antibody levels between six and twelve month sampling (n=29), late registration of vaccination records (n=2) or overlapping six and twelve month sampling (n=2). Therefore, the subset from which we further assessed the persistence of neutralizing and IgG antibodies a year after SARS-CoV-2 infection consisted of 367 subjects. Participant demographics and clinical characteristics for the selected cohort were similar to the overall cohort and are shown in Table 1. NAb, S-IgG and N-IgG antibodies were detected in 91%, 98% and 67% of subjects in the selected cohort at six months after infection, respectively (Table 2). One year after infection the proportion of positive samples was still high for NAb and S-IgG (89% (326/367) and 97% (356/367)), respectively, but had decreased to 36% (132/367) for N-IgG. The mean IgG concentrations decreased significantly (p<0.001) for SFL-IgG, RBD-IgG and N-IgG from six months (3.2, 2.3, 1.2 binding antibody units (BAU)/ml) to twelve months (2.3, 1.7, 0.44 BAU/ml, respectively) after infection. The decrease in mean IgG concentration was more notable (2.7-fold) for N-IgG compared to SFL-IgG or RBD-IgG (1.4-fold) (Figure 2).

**Table 2.** Number and proportion of positive samples for spike protein IgG (S-IgG) and neutralizing antibodies (NAb) by disease severity, age and gender of the participants six and twelve months after infection, n=367

| Disease severity | Age (years) | Gender | S-IgG positive<br>n/n (%) |  |  |  | NAb positive<br>(wt) n/n (%) |  |  |  |
| --- | --- | --- | --- | --- | --- | --- | --- | --- | --- | --- |
|  |  |  | Six months |  | Twelve months |  | Six months |  | Twelve months |  |
| Severe | ≥60 | M | 16/16 | (100) | 16/16 | (100) | 16/16 | (100) | 16/16 | (100) |
|  |  | F | 18/18 | (100) | 18/18 | (100) | 17/18 | (94) | 18/18 | (100) |
|  | >60 | M | 6/6 | (100) | 6/6 | (100) | 6/6 | (100) | 6/6 | (100) |
|  |  | F | 7/7 | (100) | 7/7 | (100) | 7/7 | (100) | 7/7 | (100) |
| Mild | ≥60 | M | 120/122 | (98) | 117/122 | (96) | 105/122 | (86) | 99/118 | (84) |
|  |  | F | 166/171 | (97) | 165/171 | (97) | 159/171 | (93) | 151/171 | (88) |
|  | >60 | M | 15/15 | (100) | 15/15 | (100) | 15/15 | (100) | 14/15 | (93) |
|  |  | F | 12/12 | (100) | 12/12 | (100) | 10/12 | (83) | 12/12 | (100) |

**Figure 2.**
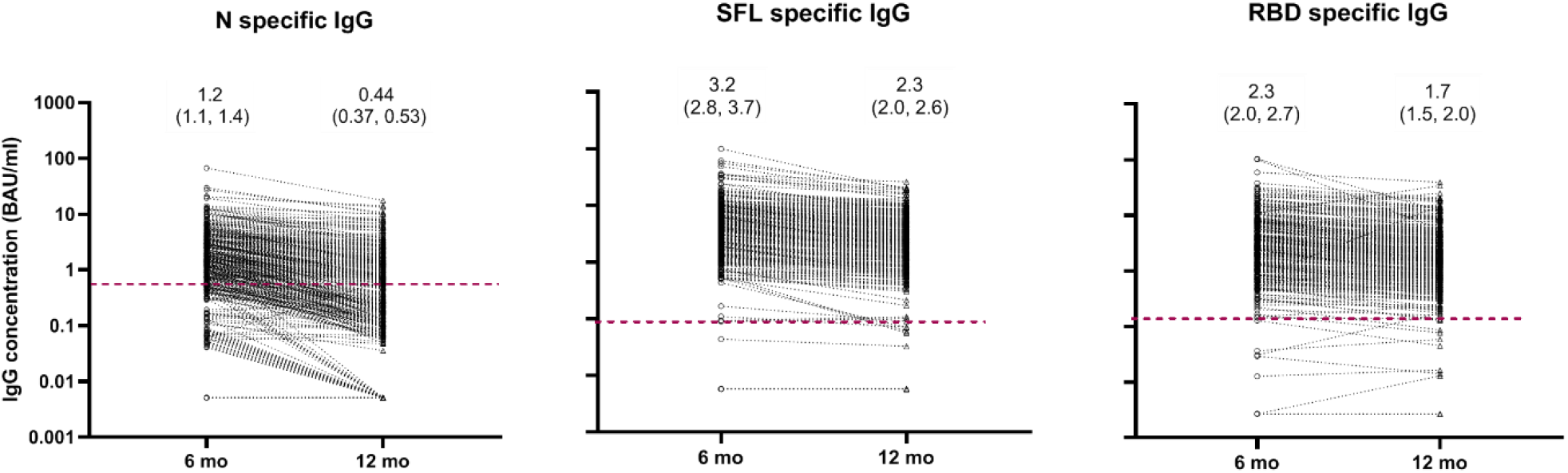
Nucleoprotein (N), and spike protein (SFL, RBD) specific IgG concentrations (BAU/ml) with geometric mean concentrations (95% CI) at six and twelve months after infection, n=367. FMIA specific cut-off for seropositivity is indicated by a dashed red line.

### Effect of disease severity, age and gender on SARS-CoV-2 antibodies

We observed higher mean N-IgG, SFL-IgG and IgG-RBD concentrations in subjects who had recovered from severe disease than in those with mild disease six months after infection (p<0.001; Figure 3). The difference was 2.0 to 7.4-fold, depending on the age group, and persisted for at least twelve months after infection (Figure 3, Table 3). The proportion of seropositive subjects with severe disease remained high for S-IgG and NAb (100%) and relatively high for N-IgG (67%) a year after severe infection, compared to 97%, 87% and 32%, respectively, of those with a milder infection. A higher proportion (33%) of subjects in the elderly age group (≥60 years of age) had been hospitalized compared to the younger age groups (13% of 40 to 59 years and 6% of those 17 to 39 years of age). Elderly subjects (≥60 years of age) with mild infection had similar levels of S-IgG antibodies and equally high proportion of the elderly subjects had NAb compared to younger subjects. N-IgG concentrations were, however, higher among ≥60-year old subjects than in subjects <60 years of age with mild disease at six and twelve months after infection (p<0.01). We could not demonstrate any difference in N-, SFL-or RBD-IgG concentrations between males and females at six or twelve months after infection.

**Figure 3.**
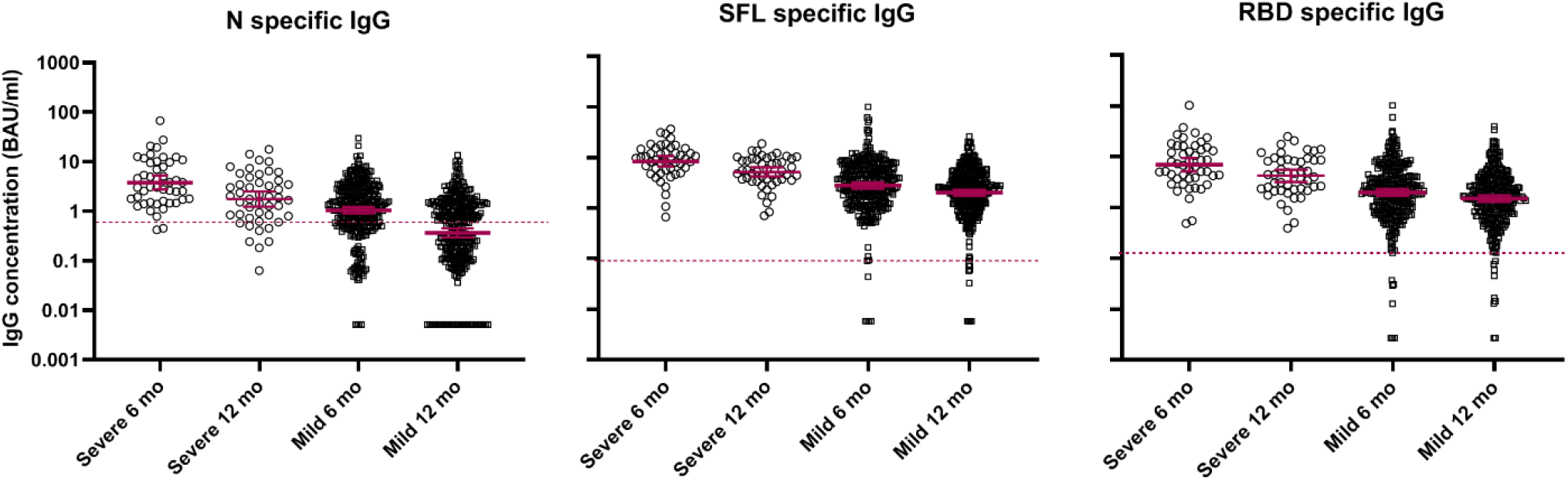
Distribution and geometric mean of IgG concentrations (BAU/ml and 95% CIs) for nucleoprotein (N specific IgG), and spike protein (SFL and RBD specific IgG) in subjects six and twelve months after severe (n=47) or mild (n=320) infection. FMIA specific cut-off for seropositivity is indicated by a dashed red line.

**Table 3.** Geometric mean IgG concentrations, GMC [95% CI], expressed as BAU/ml for nucleoprotein (N), spike proteins (SFL and RBD) at six and twelve months after COVID-19 infection per age group and disease severity. Significantly higher (Kruskal-Wallis test, p<0.05) IgG concentrations in subjects with severe as compared to mild disease within age groups are shown in bold.

| Disease | Age (years)<br>n | N-IgG<br>GMC [95% CI] |  | RBD-IgG<br>GMC [95% CI] |  | SFL-IgG<br>GMC [95% CI] |  |
| --- | --- | --- | --- | --- | --- | --- | --- |
|  |  | Six months | Twelve months | Six months | Twelve months | Six months | Twelve months |
| Mild | **17-39**<br>n=101 | 0.71<br>[0.40–1.0] | 0.23<br>[0.0061–0.46] | 1.7<br>[0.87–2.5] | 1.5<br>[0.85–2.1] | 2.5<br>[1.1–4.0] | 2.0<br>[1.4–2.6] |
|  | **40-59**<br>n=192 | 1.2<br>[0.74–1.6] | 0.41<br>[0.18–0.64] | 2.0<br>[0.68–3.4] | 1.5<br>[0.83–2.1] | 2.8<br>[1.4–4.1] | 1.9<br>[1.4–2.5] |
|  | **≥60**<br>n=27 | 2.1<br>[0.32–3.9] | 0.81<br>[-0.29–1.9] | 3.0<br>[1.1–4.8] | 1.9<br>[-0.70–4.5] | 4.0<br>[1.9–6.1] | 2.6<br>[1.1–4.2] |
| Severe | **17-39**<br>n=6 | 2.9<br>[-1.9–7.7] | **1.7**<br>**[-0.33–3.7]** | 8.4<br>[-33–50] | 4.6<br>[0.19–9.0] | 6.9<br>[0.35–14] | 5.1<br>[0.82–9.4] |
|  | **40-59**<br>n=28 | **3.9**<br>**[-0.96–8.7]** | **1.8**<br>**[0.29–3.2]** | **6.2**<br>**[3.5–8.8]** | **4.0**<br>**[1.8–6.2]** | **7.6**<br>**[5.2–10]** | **4.7**<br>**[3.2–6.3]** |
|  | **≥60**<br>n=13 | 4.1<br>[-1.4–9.5] | 1.8<br>[-0.91–4.5] | 8.4<br>[1.4–15] | 4.5<br>[0.89–8.1] | **11.4**<br>**[5.2–18]** | 6.4<br>[4.1–8.7] |

### Antibody responses indicating vaccination or reinfection

Since our focus was to investigate persistence of humoral immunity after infection, 29 (7.3%) individuals were excluded from the analysis on the basis of ≥30% increase in SFL-IgG and RBD-IgG antibody concentrations between six and twelve months after the index infection and with no documentation of received SARS-CoV-2 vaccination. Of these 29 individuals, five had strongly increased (19 to 342-fold) SFL- and RBD-IgG levels without any increase in N-IgG levels. This suggests that they may have received SARS-CoV-2 vaccination which was not documented in the National Vaccination Registry. Additionally, 15 of these 29 individuals had notable (1.3 to 3.3-fold) increases measured against both S-IgG antigens but not to N-IgG. Interestingly, nine of these 29 individuals had ≥30% increases in both S-IgG and N-IgG concentrations suggesting they may have had a re-infection with SARS-CoV-2. All 29 individuals had NAb at six and twelve months after infection. None of the 29 subjects excluded from the main analysis due to increases in S-IgG concentrations had a documented PCR-confirmed re-infection in the National Infectious Diseases Register.

### Comparison of neutralizing antibody (NAb) titers between a wild-type virus and three variants of concern (VOC)

A smaller age- and gender-matched subset of participants (n=78) was randomly selected to NAb titration of twelve-month samples due to the laborious live-virus microneutralization (MNT) assay requiring biosafety level 3 facility. The samples were re-analyzed against a wild-type virus isolated in Finland during 2020 (Fin1-20) and variants of concern (VOC) isolated in Finland during 2021: Fin34-21, Fin32-21 and Fin37-21 standing for Alpha, Beta and Delta variant, respectively. The samples to be included in the NAb titration were initially selected based on a seropositive result (NAb titer ≥6) in the screening test. The majority of the samples remained positive in the wild-type virus reanalysis, but one subject within mild disease category had a low positive result (borderline) (Figure 4, supplementary Table 1), which reflects the variability of the biological test.

**Figure 4.**
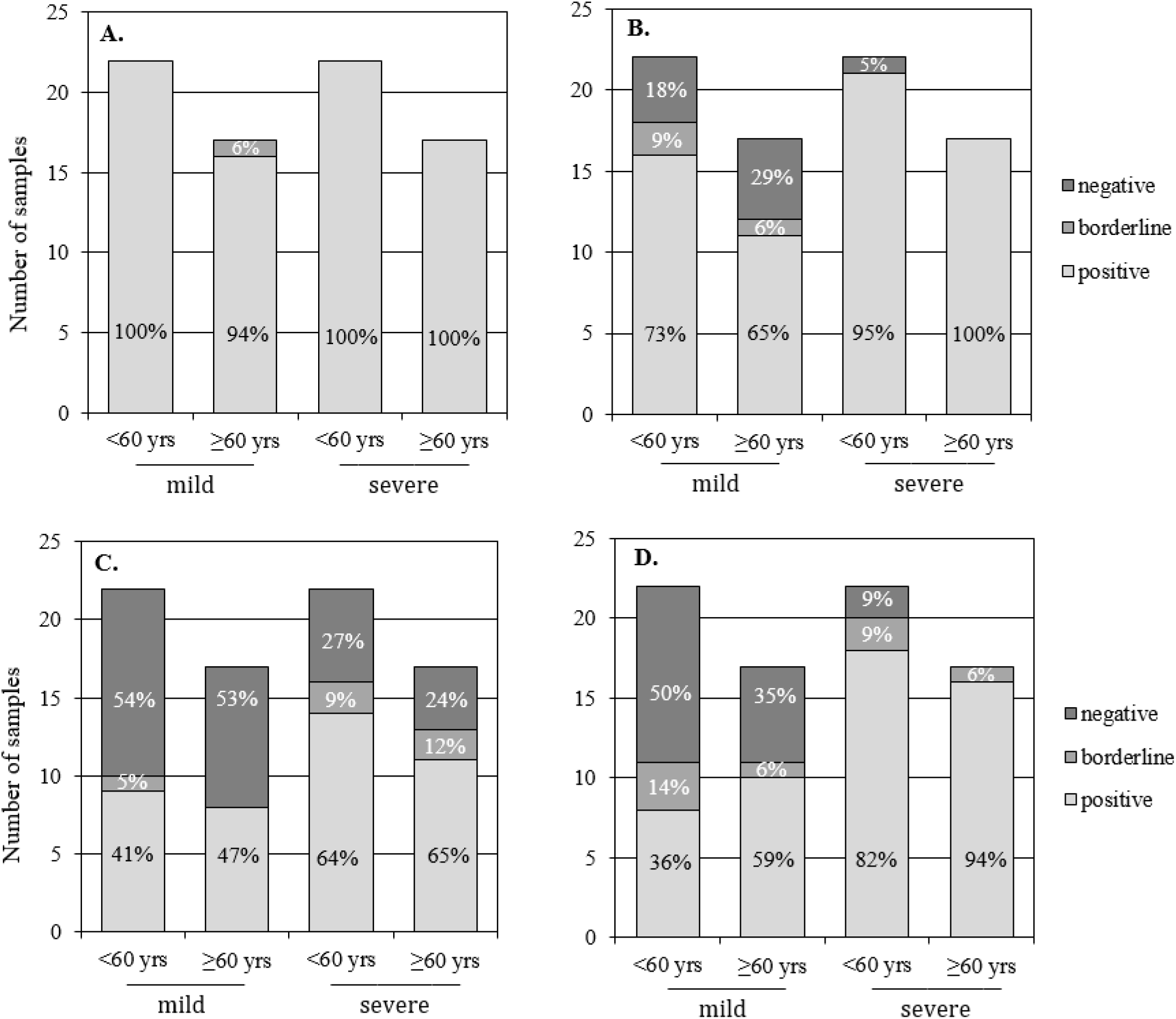
The proportion of subjects positive, low positive (borderline) and negative for neutralizing antibodies twelve months after infection against four SARS-CoV-2 virus strains (n=78). **A**. Wild-type virus (B.1). **B**. Alpha variant (B.1.1.7). **C**. Beta variant (B.1.351). **D**. Delta variant (B.1.617.2).

Within the whole cohort (n=78), NAb titers were significantly lower for all VOCs (p<0.0001, supplementary Table 2) compared to wild-type virus titers; 1.2 to 2.2-fold, 3.3 to 6.6-fold and 2.6 to 3.5-fold lower geometric mean titers (GMT) were observed for Alpha, Beta and Delta variant, respectively (Table 4). NAb titers for all VOCs correlated well with wild-type virus titers, yet more pronounced correlation was seen for Alpha and Delta variants and lower for Beta variant (supplementary Figure 1)

**Table 4.** Geometric mean IgG concentrations, GMC [95% CI] expressed as BAU/ml for nucleoprotein (N) and spike proteins (SFL and RBD) and geometric mean titers, GMT [95% CI] of neutralizing antibodies (NAb) against wild-type (wt) virus and three variants of concern Alpha (B.1.1.7), Beta (B.1.351) and Delta (B.1.617.2) twelve months after infection (n=78).

| Disease severity | Age | Gender | n | IgG concentration (BAU/ml) |  |  | MNT titer |  |  |  |
| --- | --- | --- | --- | --- | --- | --- | --- | --- | --- | --- |
|  |  |  |  | N-IgG | S-IgG (RBD) | S-IgG (SFL) | NAb wt | NAb Alpha | NAb Beta | NAb Delta |
| Severe | <60y | M+F | 22 | 1.5<br>[0.88-2.7] | 3.9<br>[2.5-6.1] | 4.7<br>[3.0-7.2] | 27<br>[17-41] | 21<br>[14-34] | 8.1<br>[5.0-13] | 10<br>[7.1-15] |
|  |  | M | 12 | 2.0<br>[0.99-4.0] | 4.7<br>[2.4-9.0] | 5.5<br>[2.9-10.4] | 29<br>[16-55] | 26<br>[14-49] | 9.2<br>[4.6-19] | 14<br>[8.1-23] |
|  |  | F | 10 | 1.1<br>[0.45-2.9] | 3.2<br>[1.8-5.7] | 3.8<br>[2.1-6.8] | 24<br>[12-47] | 17<br>[8.5-32] | 6.8<br>[3.6-13] | 7.7<br>[4.5-13] |
|  | ≥60y | M+F | 17 | 1.6<br>[0.98-2.5] | 5.1<br>[3.0-8.7] | 7.6<br>[4.8-12] | 52<br>[39-71] | 30<br>[20-44] | 8.0<br>[5.1-13] | 15<br>[10-22] |
|  |  | M | 8 | 0.89<br>[0.60-1.3] | 4.2<br>[2.2-8.0] | 5.8<br>[3.6-9.2] | 39<br>[27-57] | 28<br>[18-42] | 9.2<br>[4.8-18] | 13<br>[8.5-21] |
|  |  | F | 9 | 2.6<br>[1.3-5.0] | 6.1<br>[2.6-14] | 9.7<br>[4.6-21] | 68<br>[45-100] | 32<br>[16-61] | 7.0<br>[3.6-14] | 16<br>[8.6-31] |
| Mild | <60y | M+F | 22 | 0.41<br>[0.22-0.75] | 1.6<br>[1.3-2.1] | 2.3<br>[1.9-2.9] | 15<br>[12-20] | 8.0<br>[5.4-12] | 3.6<br>[2.7-4.8] | 4.0<br>[2.8-5.7] |
|  |  | M | 12 | 0.36<br>[0.14-0.93] | 1.3<br>[0.89-1.8] | 1.8<br>[1.4-2.4] | 12<br>[9.5-16] | 5.1<br>[3.1-8.4] | 2.9<br>[2.1-4.1] | 2.9<br>[2.0-4.0] |
|  |  | F | 10 | 0.47<br>[0.22-1.0] | 2.2<br>[1.6-3.0] | 3.1<br>[2.3-4.0] | 20<br>[14-30] | 13<br>[8.3-22] | 4.6<br>[2.9-7.3] | 6.0<br>[3.4-11] |
|  | ≥60y | M+F | 17 | 0.50<br>[0.26-1.1] | 1.8<br>[1.0-3.1] | 2.1<br>[1.3-3.4] | 19<br>[11-31] | 8.5<br>[4.8-15] | 4.2<br>[2.8-6.5] | 5.6<br>[3.5-8.8] |
|  |  | M | 8 | 0.94<br>[0.42-2.1] | 1.5<br>[0.72-3.2] | 1.5<br>[0.82-2.8] | 12<br>[6.1-23] | 4.6<br>[2.2-9.7] | 2.9<br>[1.8-4.6] | 4.1<br>[2.2-7.6] |
|  |  | F | 9 | 0.28<br>[0.081-0.98] | 2.1<br>[0.91-4.7] | 2.9<br>[1.5-5.7] | 28<br>[14-55] | 15<br>[7.1-30] | 6.0<br>[3.2-11] | 7.4<br>[3.8-14] |

For both wild-type and Alpha variant the proportion of seropositive individuals with severe disease remained high twelve months after infection (Figure 4, supplementary Table 1). Lower titers against Alpha variant compared to wild-type virus were seen in mild disease groups with increasing proportion of low positive (borderline) or negative subjects. The greatest decrease of NAb titers was seen between the wild-type virus and the Beta variant with markedly lower GMTs and seropositivity with several borderline titers also in groups of severe disease. NAb titers and seropositivity for the Delta variant was also markedly lower compared to wild-type virus. The Delta GMT values placed between the GMTs of Alpha and Beta variants, yet the seropositivity of severe disease groups was relatively well preserved (≥80%) compared to that of the Beta variant (65%).

For all viruses, the subjects recovered from severe disease had overall 2.1 to 3.0-fold NAb titers compared to those with mild disease (p<0.01, supplementary Table 2). The same finding was seen with all IgG concentrations. The difference in IgG concentrations between severe and mild disease was prominent in both sexes in the large study cohort (n=367). However, in the small cohort (n=78) only males with mild disease had markedly lower NAb titers and S-IgG concentrations compared to those recovered from severe disease (p<0.05, supplementary Table 3). The difference was not statistically significant for females although the trend was similar.

NAb titers against wild-type virus were higher in the elderly group (≥60 years) compared to <60 years-old (p=0.045) whereas NAb titers for VOCs did not differ significantly between age groups (supplementary Table 4). We detected strong and statistically significant correlation (p<0.0001) between NAb titers and S-IgG antibody concentrations indicating overall parallel trend between severe and mild disease antibody levels (Figure 5).

**Figure 5.**
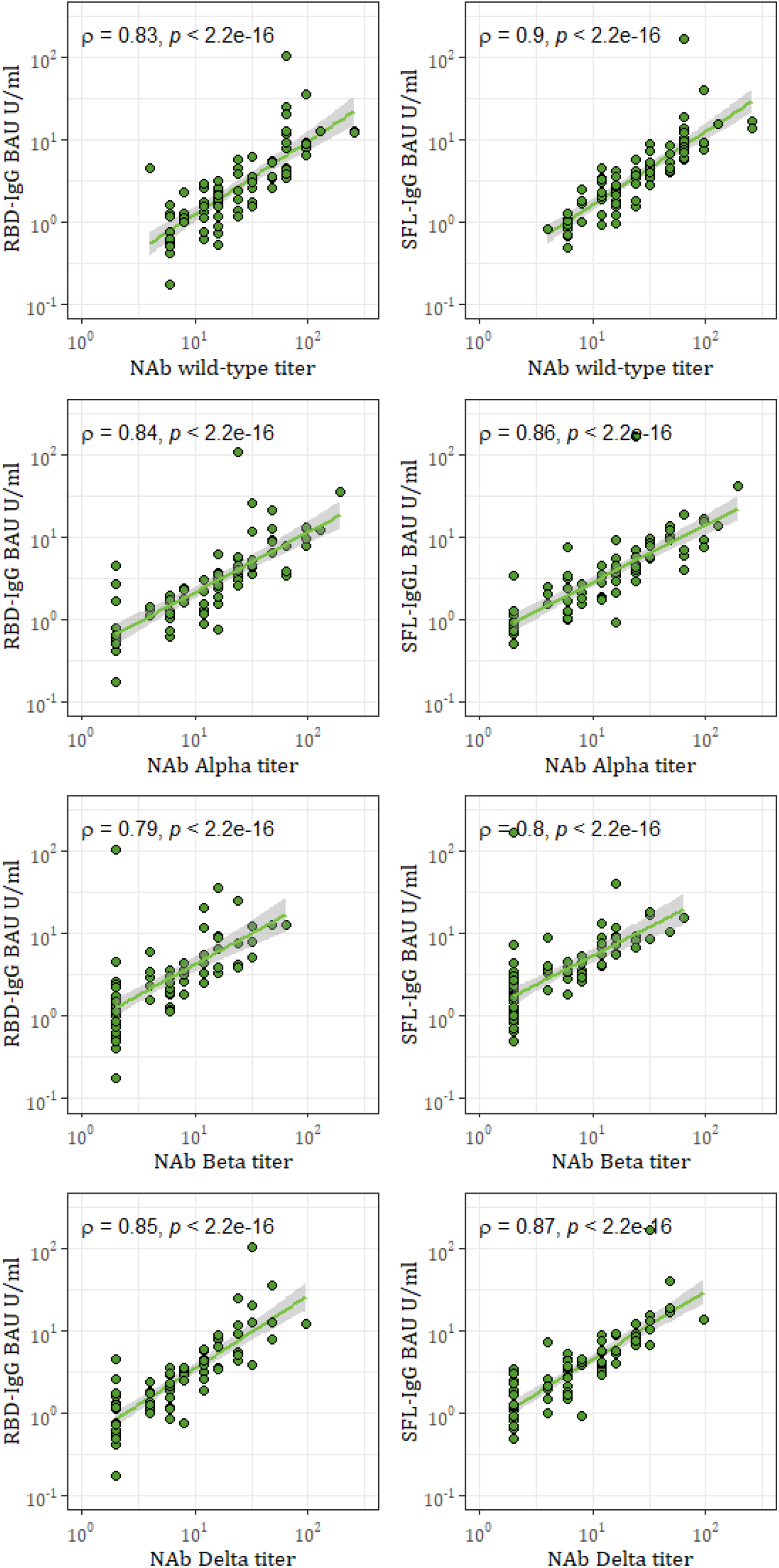
Spearman correlation (ρ) and significance (*p*) between S-IgG antibody concentrations and neutralizing antibody (NAb) titers against the wild-type virus (B.1) and variants of concern: Alpha (B.1.1.7), Beta (B.1.351) and Delta (B.1.617.2). One point may represent multiple samples (n=78).

## Discussion

Studies of individuals who have recovered from natural SARS-CoV-2 infection are crucial in determining how long antibodies persist after infection and whether these antibodies might protect against re-infection. We showed that S-IgG antibodies and, most importantly, NAbs persist in most subjects for at least a year following SARS-CoV-2 infection. The concentration of N-IgG, on the contrary, declines to a level that is not distinguishable from unspecific, cross-reactive antibodies among a large proportion of subjects. In accordance with previous observations (5, 7, 25), subjects with severe infection had higher N-IgG, S-IgG concentrations and NAb titers than subjects with mild infection and are expected to remain seropositive for longer time.

Previous studies show that most patients recovering from COVID-19 have detectable antibody responses peaking at approximately one month after infection (6, 7, 26); Dub et al. manuscript in preparation. Thereafter antibody levels to nucleoprotein and spike protein antigens decline in the first few months with differences in isotype and antigen specificity of the antibody (6). The decay rate has been shown to slow down thereafter (11). The relatively rapid early decline in S-IgG antibodies followed by slower decay indicates a transition of serum antibodies from being produced by short-lived plasmablasts to a more persistent population of long-lived plasma cells generated later in the immune response (27). Consistently, NAbs and T cell immunity have been reported to persist at least six to twelve months after infection (5-7, 10-12, 28). Our data is consistent with previous data suggesting that, even though NAb titers decline with time, NAbs persist in most subjects, at least up to a year.

We observed that markedly lower proportion of subjects had N-IgG than S-IgG antibodies at six months after infection. Thereafter the concentration N-IgG antibodies declined to a level that was not distinguishable from unspecific, cross-reactive antibodies among a large proportion of subjects twelve months after infection. SARS-CoV-2 nucleoprotein is produced abundantly during infection and since it is not a component in present vaccines or vaccine candidates it could potentially serve as a measure of past infection. However, our results clearly show that as time after infection increases, the sensitivity of our N-IgG based antibody assay decreases. In agreement to our findings the more rapid decay of N-IgG after SARS-CoV and SARS-CoV-2 infection has also been reported in another studies (26, 29). In contrast to our results, some studies have reported up to 100% of patients having N-IgG nearly a year after infection (8) or comparable persistence to spike protein specific IgG antibodies (6). Different assay platforms and cut-offs for N-IgG seropositivity may explain the observed differences. The cut-off used in this study (0.58 BAU/ml) was based on 98.6% sensitivity and 100% specificity of the in-house FMIA assay for samples taken 13 to 150 days post onset of symptoms and is directly bridged to the WHO International SARS-CoV-2 Standard (NIBSC code 20/136) (30); Solastie et al. manuscript in preparation.

Even though NAbs persist relatively long in most subjects, neutralization efficiency against Alpha (B.1.1.7), Beta (B.1.351) and Delta (B.1.617.2) variants was decreased compared to the wild-type virus. This was emphasized in subjects who had recovered from mild disease which is concerning since mild symptomatic individuals represent the majority of COVID-19 cases (1). Indeed, NAbs may not be developed in mild symptomatic or asymptomatic individuals and may also wane relatively quickly after infection (31).

In line with earlier observations nine (12) and twelve months after infection (11), we found that NAb levels against the Alpha variant had only slightly, although statistically significantly reduced, while NAb levels against the Beta variant were considerably declined compared to the wild-type virus. Beta variants have shown to evade antibody responses induced upon infection as well as vaccination (15-17, 32, 33). Although the NAb levels were declined against the Beta variant, we observed that over 60% of hospitalized subjects were seropositive a year after infection, which indicates long-lived cross-neutralization capacity induced by severe disease.

We detected substantially declined NAb titers against Delta variant in subjects with mild disease, similar to what has been previously reported after vaccination or from 5-20 weeks to up to 12 months after SARS-CoV-2 infection (18, 19, 34-36). However, we observed that over 80% of the subjects who had recovered from severe disease were seropositive against the Delta variant. This is in line with one study reporting only modestly reduced (88%) NAb levels against Delta variant 2-4 weeks after second vaccine dose when antibody concentrations are peaking (37). Our results support the previous findings that emerging variant Delta partially but significantly escapes NAbs (18, 19).

One previous study reported lower seropositivity rates one year after mild SARS-CoV-2-infection compared to our results; 58% were positive for S1-IgG and 85% for S-IgG measured with enzyme immunoassay and 58% had NAb (10). Direct comparison of the IgG concentrations and NAb titers between studies may not be possible since the age groups, viruses, as well as the serological tests, differed. Neutralizing antibody tests have not been standardized and among other things the starting dilutions of serum samples may vary between assays. The microneutralization assay used in this study utilized live virus and the starting dilution of 1:4 further enhances the sensitivity of the assay in detecting low levels of NAbs.

In our study population, we could not see a gender effect in hospitalized individuals, as previously reported (5, 25). However, hospitalized subjects ≥60 years tended to have slightly higher IgG and NAb levels compared to hospitalized subjects <60 years suggesting more severe infection in the elderly age group. Although there was no overall difference between the genders, especially males with mild disease had markedly lower NAb titers for all viruses compared to individuals who recovered from severe disease.

An interesting subgroup within our study consisted of 29 out of 400 selected individuals with notable (≥30%) increase in S-IgG antibody concentrations from six to twelve months after infection with no documentation of immunization with any COVID-19 vaccines. Increasing N- and S-IgG antibodies in 9 of these individuals suggest they have re-encountered SARS-CoV-2 and have been able to mount a memory IgG response to SARS-CoV-2 that may be detectable even months after the encounter. Taking into consideration the more rapid decline of N-IgG than S-IgG antibodies described in this study, also the responses of the 15 individuals with an increase in solely S-IgG concentrations may indicate re-infection. None of these individuals had documentation of PCR-confirmed re-infection in the National Infectious Diseases Register, which may indicate asymptomatic or mild infection.

There is a major research and development effort to produce effective SARS-CoV-2 vaccines. The long-term kinetics and persistence of immunity after vaccination is, however, largely unknown. Evidence from convalescent sera from individuals who have recovered from natural infection may help determine how long antibodies and immunity persist, and whether antibodies might protect against re-infection. Previous data shows that, when measured as IgG antibodies against spike protein or RBD and NAb, immune response after two doses of SARS-CoV-2 vaccine is similar to that observed in convalescent sera from COVID-19 patients (38-41). Evidence of persistence of antibodies and immunity after infection will help in predicting persistence of immunity after SARS-CoV-2 vaccination.

We recognize certain limitations in our study. Due to high SARS-CoV-2 vaccine coverage in the older age groups (≥60 years of age) at the time of the study, only 11% of the participants of our study were ≥60 years of age, the age group with highest disease incidence and morbidity. Since the median age of study subjects was 50 years, our results may not necessarily apply to all age groups. The number of subjects selected to the NAb titer comparison was limited but the study subjects were matched by disease severity, age and gender and randomly selected from the participants.

Previous studies have indicated that the presence of antibodies to SARS-CoV-2 was associated with a significantly reduced risk of SARS-CoV-2 reinfection among health-care workers for up to seven months after infection (21, 42). We observed that S-IgG antibodies and neutralizing antibodies persist at least a year after infection in the vast majority of individuals who have recovered from infection. This strongly suggests that protection against re-infection is long-lived, although antibody-mediated immunity may not persist equally well among elderly subjects. A previous study found that patients older than 60 years had fewer memory B cells secreting total IgG and RBD-specific IgG than patients under 60 years old nine months after infection (8). In this study we observed that IgG concentrations declined from six to twelve months more substantially in subjects ≥60 years compared to younger age groups. Similar more rapid decline in neutralizing antibody concentrations was observed among elderly compared to younger subjects who were followed up to six months following vaccination (43). The results of our study support previous findings indicating that protection against infection mediated by neutralizing antibodies may be impaired against the variants of concern, especially after a mild disease. While in the absence of neutralizing antibodies reinfection is possible, cellular immunity is not similarly affected by mutations in the RBD site (16) and is likely to provide long-term protection against severe disease.

## Methods

### Study design and participants

In October 2020, 2586 subjects ≥18 years of age, whose native language was Finnish or Swedish and who were living within five selected hospital districts in Finland and with a PCR-confirmed COVID-19 diagnosis between February and April 2020 were identified in the National Infectious Disease Register and invited by mail to participate in the follow-up study for the serological population study of the coronavirus epidemic. Subjects within institutional care or who had participated in other serological population studies of SARS-CoV-2 conducted by the Finnish Institute for Health and Welfare were excluded from the study. The study protocol was reviewed and approved by the ethical committee the Hospital District of Helsinki and Uusimaa (HUS) and registered under the Development of seroprevalence in Finland during the new coronavirus (SARS-CoV-2) epidemic – serological population study protocol HUS/1137/2020. Informed consent was obtained from all study subjects before sample collection. A total of 1292 subjects (median age 50.0, range 17.3-94.3) with PCR-confirmed COVID-19 participated and donated a blood sample for determination of SARS-CoV-2 specific serum antibodies 5.9 to 9.9 months (median 7.6 months) after infection. All those previously enrolled and still living in the same hospital district (n=1227) were invited to a follow-up visit and blood sampling a year after the COVID-19 diagnosis in March-April 2021. By May 21^st^ 2021 altogether 995 participants (median age 52.5, range 17.6-95.6 years) had participated and donated a blood sample for determination of SARS-CoV-2 specific serum antibodies at 12.7 months (median, range 11.7 to 14.3 months) after the diagnosis of PCR-confirmed COVID-19. Demographics, clinical characteristics and SARS-CoV-2 vaccination history of the participants were collected from the National Infectious Diseases Register, the Care Register for Health Care and the Register of Primary Health Care Visits and the National Vaccination Registry and are summarized in Table 1. Since late December 2020 SARS-CoV-2 vaccinations have been offered according to the national recommendations in Finland. In this study the disease severity was defined as severe or mild. Severe infection was defined as an individual with laboratory confirmed COVID-19 and who required hospital treatment. Mild infection was defined as an individual with laboratory confirmed COVID-19 without hospital treatment.

### Sample processing and selection of samples

Sera were separated by centrifugation, aliquoted and stored at -20°C or below. For assessment of NAbs, sera were heat-inactivated (56°C for 30 min) and then stored at -20°C or below.

For assessment of persistence of serum antibodies six months following PCR-confirmed COVID-19 diagnosis, all samples taken ≤10 months after diagnosis (n=1292) were selected for assessment of SARS-CoV-2 IgG antibody concentration and neutralizing antibodies (positive/borderline/negative). For assessment of antibody persistence twelve months after infection, 400/995 sera were randomly selected for determination of SARS-CoV-2 IgG antibody concentration and neutralizing antibodies. Selection criteria were: 6-month sample available, PCR-confirmed COVID-19 diagnosis, no documentation of SARS-CoV-2 vaccination in the Register of Primary Health Care Visits by June 10^th^ 2021. Further, samples of subjects with ≥30% increase in IgG antibody concentration to both SARS-CoV-2 spike glycoprotein antigens (full-length spike protein (SFL) and receptor binding protein (RBD)) between six and twelve month blood sampling (n=29) were excluded from the analysis. An additional four samples were excluded due to late discovery of these samples not meeting selection criteria. Of the four samples two were excluded due to vaccination and two due to samples taken over 10 months after infection. Consequently, 367 sera were selected.

### A smaller subset of participants for the NAb titration

For comparison of NAb titers against a wild-type virus and variants of concern (Alpha, Beta and Delta), 80/536 twelve-month sera screened to have NAb positive (titer ≥6 against wild-type virus) were randomly selected as above mentioned. Later observed ≥30% increase in IgG antibody concentration between six- and twelve-month samples excluded 2/80 samples leaving total sample size to 78. SARS-CoV-2 IgG antibody concentration was measured from this cohort to ensure its comparability to the other 367 sera selected.

### SARS-CoV-2 microneutralization test (MNT)

A cytopathic effect (CPE) based microneutralization test (MNT) was performed as previously described (30, 44). Briefly, heat-inactivated serum samples were 2-fold serially diluted starting from 1:4 in EMEM supplemented with penicillin, streptomycin and 2% of heat-inactivated Fetal Bovine Serum (FBS). At the BSL-3 laboratory, pre-titrated virus was added to obtain 100 TCID_50_ per well following incubation for 1 h at +37°C, 5% CO_2_. African green monkey kidney epithelial (VeroE6) cells were added and the 96-well tissue culture plates were incubated at +37°C, 5% CO_2_ for 4 days. Wells were fixed with 30% formaldehyde and stained with crystal violet. Results were expressed as MNT titers corresponding to reciprocal of serum dilution that inhibited 50% of SARS-CoV-2 infection observed by CPE of inoculated cells. MNT titer ≥6 was considered positive, borderline when 4 and negative when <4. Borderline values were further confirmed with biological repeats. For titer comparison, a titer of 192 was measured for the NIBSC-standard 20/136 (45) using the wild-type virus Fin1-20.

### SARS-CoV-2 viruses selected for MNT

All samples were screened with wild-type virus Fin1-20 (B.1 lineage): hCoV-19/Finland/1/2020 (GISAID accession ID EPI_ISL_407079) for NAb positivity. Fin1-20 was the first SARS-CoV-2 strain detected in Finland in January 2020. Virus isolation and propagation was performed in Vero E6 cells (44). A smaller subset of samples was analyzed also with variants of concern (VOC) isolated in Finland during 2021: Fin34-21, Fin32-21 and Fin37-21 which stand for Alpha, Beta and Delta variant, respectively. Alpha variant (B.1.1.7) Fin34-21 indicates the isolate hCoV-19/Finland/THL-202102301/2021 (EPI_ISL_2590786). The virus was detected in January 2021. Fin32-21 as a representative of Beta variant (B.1.351) was detected in January 2021. Spike region sequenced from the patient sample showed typical Beta variant amino acid changes. Delta variant (B.1.617.2) Fin37-21 was detected in May 2021 and indicates hCoV-19/Finland/THL-202117309/2021 (EPI_ISL_2557176). All variant viruses were isolated and propagated (passages 1-2) in VeroE6-TMPRSS2-H10 cells (46) and further propagated in Vero E6 cells (passage 3) for MNT.

### SARS-CoV-2 fluorescent microsphere immunoassay (FMIA)

The SARS-CoV-2 FMIA has been previously described in detail by Ekström et al. (30) and Solastie et al. (manuscript in preparation). Briefly, diluted sera, reference and controls were added onto 96-well plates with MagPlex®-C superparamagnetic carboxylated microspheres (Luminex Corp) conjugated with SARS-CoV-2 nucleoprotein (N, REC31812), full-length spike protein (SFL, REC31868) and receptor binding domain of spike protein (RBD, REC31849, The Native Antigen Company). Antigen covered microspheres were let to incubate with sera for 1 h, after which unbound particles were washed away with a magnetic plate washer (ELx405 and 405TSRS, BioTek). R-Phycoerythrin-conjugated Affinipure Goat Anti-Human IgG detection antibody (Jackson Immuno Research) was added into the wells and incubated for 30 minutes before washing. Median fluorescence intensity (MFI) was measured with MAGPIX system (Luminex) and binding antibody unit concentrations (BAU U/ml) were interpolated from 5-parameter logistic curves with xPONENT (v. 4.2, Luminex) created by 7-point serial 4-fold diluted reference sera calibrated against WHO International Standard (NIBSC code 20/136; (45); Solastie et al. manuscript in preparation). Serum samples were analysed as true duplicates in 1:100 and 1:1600 dilutions. If antibody concentration exceeded the linear range of the reference the sample was re-analysed with higher dilution. When MFI of a sample was below the linear range of the reference, the sample was assigned an antibody concentration half of the limit of detection (0.0094, 0.012 and 0.0057 BAU/ml for N-, SFL- and RBD-IgG). A sample was considered positive for SARS-CoV-2 IgG antibodies (S-IgG) when SFL and RBD specific antibody concentrations were ≥0.089 and ≥0.13 BAU/ml, respectively. A sample was considered positive for nucleoprotein specific IgG antibodies (N-IgG) when N-IgG concentration was ≥0.58 BAU/ml. The cut-offs for seropositivity were determined during clinical validation of the FMIA and yielded both sensitivity and specificity of 100% for SFL- and RBD-IgG and 98.6% and 100% for N-IgG for samples taken 13 to 150 days post onset of symptoms, respectively (30); Solastie et al. manuscript in preparation.

### Statistical methods

We calculated the geometric mean concentrations (GMC) and geometric mean titers (GMT) with 95% confidence intervals (CI) for IgG and neutralizing antibodies, respectively. We assessed the statistical differences in antibody levels between groups using Kruskal-Wallis test with Bonferroni correction. Differences in mean IgG concentrations between six and twelve months after infection were compared using Students paired t-test and log-transformed data. The statistical significance level of difference was set to p<0.05. We used Spearman correlation in the determination of correlation coefficients and significance. MNT titers <4 were assigned a titer value of 2. Samples with IgG concentrations below the limit of detection were assigned an antibody concentration equal to half of the limit of detection. Statistical analyses were performed using SPSS v27 and R (v4.0.4) with Rstudio (v1.4.1106).

## Data Availability

The data that support the findings of this study are available from the corresponding author upon reasonable request. The SARS-CoV-2 sequence data used in this study have been deposited in the GISAID EpiCoV™ Database under isolate codes EPI_ISL_407079, EPI_ISL_2590786 and EPI_ISL_2557176.

## Data availability statement

The data that support the findings of this study are available from the corresponding author upon reasonable request. The SARS-CoV-2 sequence data used in this study have been deposited in the GISAID’s EpiCoV™ Database under isolate codes EPI_ISL_407079, EPI_ISL_2590786 and EPI_ISL_2557176.

## Acknowledgements

We thank all the study participants. We also thank Tuomo Nieminen, Esa Ruokokoski, Juha Oksanen, Dennis Ahlfors, Timo Koskenniemi, Katja Lind, Hanna Valtonen, Heidi Hemmilä, Marja-Liisa Ollonen, Larissa Laine, Tiina Sihvonen, Raisa Hanninen, Johanna Rintamäki, Leena Saarinen, Marja Suorsa, Lotta Hagberg, Mervi Lasander, Marja Leinonen, Hanna K. Valtonen, Jenni Krogell, Arja Rytkönen, Susanna Celik, Maila Kyrölä, Päivi Siren and Timothée Dub.

We gratefully acknowledge the authors and their respective laboratories, who analyzed and submitted the sequences to GISAID’s EpiCoV™ Database.

## Author contributions

M.M., N.E. and A.H. designed the experiments. M.M., A.A.P. and HN contributed to the study design. C.V. and A.S. developed and performed the FMIA tests. A.H. developed and performed the microneutralization tests. P.Ö. coordinated the virus isolations. A.H., N.E. and A.S. analyzed the data. E.I. and N.E coordinated the participant recruitment, sample collection and sample processing. A.H., N.E. and M.M. wrote the manuscript and all co-authors contributed to the edition of the text.

