## Supplementary Figure 1 for "Persistence of neutralizing antibodies a year after SARS-CoV-2 infection"

**Supplementary Figure 1.** Spearman correlation (ρ) and significance (*p*) between neutralizing antibody (NAb) titers against the wild-type virus (B.1) and variants of concern: Alpha (B.1.1.7), Beta (B.1.351) and Delta (B.1.617.2). One point may represent multiple samples (n=78).

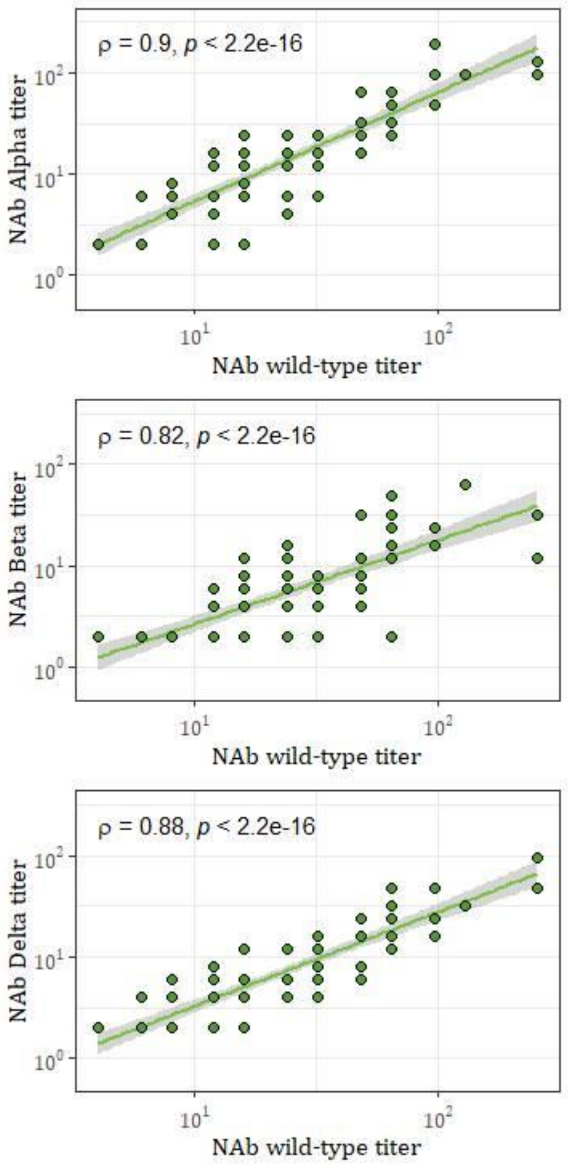
