## Supplementary Table 1 for "Persistence of neutralizing antibodies a year after SARS-CoV-2 infection"

**Supplementary Table 1.** The proportion of subjects (M=male; F=female) positive for nucleoprotein (N) and spike proteins (SFL and RBD) IgG antibodies and the proportion of subjects positive and low positive (borderline) for neutralizing antibodies (NAb) twelve months after infection against four SARS-CoV-2 virus strains (n=78).

| | | | | **IgG positivity%** | | | **MNT positivity (%)**<br>***MNT borderline (%)*** | | | |
|---|---|---|---|---|---|---|---|---|---|---|
| **Disease severity** | **Age** | **Gender** | **n** | **N-IgG** | **S-IgG (RBD)** | **S-IgG (SFL)** | **NAb wt** | **NAb Alpha** | **NAb Beta** | **NAb Delta** |
| **Severe** | <60y | M+F | 22 | 18/22 (82) | 22/22 (100) | 22/22 (100) | 22/22 (100) | 21/22 (95) | 14/22 (64)<br>*2/22 (9)* | 18/22 (82)<br>*2/22 (9)* |
| | | M | 12 | 10/12 (83) | 12/12 (100) | 12/12 (100) | 12/12 (100) | 12/12 (100) | 8/12 (67) | 10/12 (83)<br>*2/12 (17)* |
| | | F | 10 | 8/10 (80) | 10/10 (100) | 10/10 (100) | 10/10 (100) | 9/10 (90) | 6/10 (60)<br>*2/10 (20)* | 8/10 (80) |
| | ≥60v | M+F | 17 | 14/17 (82) | 17/17 (100) | 17/17 (100) | 17/17 (100) | 17/17 (100) | 11/17 (65)<br>*2/17 (12)* | 16/17 (94)<br>*1/17 (6)* |
| | | M | 8 | 6/8 (75) | 8/8 (100) | 8/8 (100) | 8/8 (100) | 8/8 (100) | 5/8 (63)<br>*2/8 (25)* | 8/8 (100) |
| | | F | 9 | 8/9 (89) | 9/9 (100) | 9/9 (100) | 9/9 (100) | 9/9 (100) | 6/9 (67) | 8/9 (89)<br>1/9 (11) |
| **Mild** | <60v | M+F | 22 | 11/22 (50) | 22/22 (100) | 22/22 (100) | 22/22 (100) | 16/22 (73)<br>*2/22 (9)* | 9/22 (41)<br>*1/22 (5)* | 8/22 (37)<br>*3/22 (14)* |
| | | M | 12 | 6/12 (50) | 12/12 (100) | 12/12 (100) | 12/12 (100) | 6/12 (50)<br>*2/12 (17)* | 3/12 (25)<br>*1/12 (8)* | 2/12 (17)<br>*2/12 (17)* |
| | | F | 10 | 5/10 (50) | 10/10 (100) | 10/10 (100) | 10/10 (100) | 10/10 (100) | 6/10 (60) | 6/10 (60)<br>*1/10 (10)* |
| | ≥60v | M+F | 17 | 9/17 (53) | 17/17 (100) | 17/17 (100) | 16/17 (94)<br>*1/17 (6)* | 11/17 (65)<br>*1/17 (6)* | 8/17 (47) | 10/17 (59)<br>*1/17 (6)* |
| | | M | 8 | 6/8 (75) | 8/8 (100) | 8/8 (100) | 7/8 (88)<br>*1/8 (13)* | 3/8 (38)<br>*1/8 (13)* | 2/8 (25) | 3/8 (38)<br>*1/8 (13)* |
| | | F | 9 | 3/9 (33) | 9/9 (100) | 9/9 (100) | 9/9 (100) | 8/9 (89) | 6/9 (67) | 7/9 (78) |
