## Supplementary Table 2 for "Persistence of neutralizing antibodies a year after SARS-CoV-2 infection"

**Supplementary Table 2.** Statistical analysis of neutralizing antibody titers against variant viruses Alpha (B.1.1.7), Beta (B.1.351) and Delta (B.1.617.2) compared to wild type (B1) virus (n=78). Kruskal-Wallis test with adjusted p-values.

| | **Alpha variant (B.1.1.7)** | **Beta variant (B.1.351)** | **Delta variant (B.1.617.2)** |
|---|---|---|---|
| **Wild-type virus (B1)** | 0.0000000280 | 0.0000000037 | 0.00000000086 |
