## Supplementary Table 3 for "Persistence of neutralizing antibodies a year after SARS-CoV-2 infection"

**Supplementary Table 3.** Influence of gender and disease severity twelve months after infection on spike protein IgG concentrations (SFL, RBD) and neutralizing antibody titers against wild-type (B.1) virus and variant viruses Alpha (B.1.1.7), Beta (B.1.351) and Delta (B.1.617.2) (n=78). Kruskal-Wallis with Bonferroni correction, adjusted p-values. P-values <0.05 are marked in bold.

| Compared groups | | Wild-type virus (B1) | Alpha variant (B.1.1.7) | Beta variant (B.1.351) | Delta variant (B.1.617.2) | IgG-SFL | IgG-RBD |
|---|---|---|---|---|---|---|---|
| Male mild | Male severe | **0.0029** | **0.000033** | **0.0013** | **0.000045** | **0.00022** | **0.0024** |
| Female mild | Female severe | 0.51 | 1.00 | 1.00 | 0.82 | 0.29 | 0.41 |
| Female mild | Male mild | 0.19 | **0.047** | 0.34 | 0.15 | 0.28 | 0.55 |
| Female severe | Male severe | 1.00 | 1.00 | 1.00 | 1.00 | 1.00 | 1.00 |
| Female severe | Male mild | **0.0006** | **0.00041** | **0.033** | **0.0011** | **0.00041** | **0.0024** |
| Female mild | Male severe | 1.00 | 0.40 | 0.48 | 0.18 | 0.22 | 0.42 |
