## Supplementary Table 4 for "Persistence of neutralizing antibodies a year after SARS-CoV-2 infection"

**Supplementary Table 4.** Influence of gender, age and disease severity twelve months after infection on spike protein IgG concentrations (SFL, RBD) and neutralizing antibody titers against wild type (B1) virus and variant viruses Alpha (B.1.1.7), Beta (B.1.351) and Delta (B.1.617.2) (n=78). Kruskal-Wallis with Bonferroni correction, adjusted p-values. P-values <0.05 are marked in bold.

| | **Wild-type virus (B1)** | **Alpha variant (B.1.1.7)** | **Beta variant (B.1.351)** | **Delta variant (B.1.617.2)** | **IgG-SFL** | **IgG-RBD** |
|---|---|---|---|---|---|---|
| **Gender** | 0.066 | 0.13 | 0.47 | 0.27 | 0.18 | 0.22 |
| **Age (<60 vs ≥60 yrs)** | **0.045** | 0.33 | 0.61 | 0.14 | 0.33 | 0.39 |
| **Disease severity** | **0.00022** | **0.000030** | **0.0011** | **0.000022** | **0.000015** | **0.00014** |
